# Palliative care in the treatment of women with breast cancer: a scoping review protocol

**DOI:** 10.1101/2022.03.21.22272710

**Authors:** Romel Jonathan Velasco-Yanez, Ana Fátima Carvalho Fernandes, Samuel Miranda Mattos, Thereza Maria Magalhães Moreira, Régia Christina Moura Barbosa Castro, Luís Carlos Lopes-Júnior

**Affiliations:** Department of Nursing, Federal University of Ceará, Fortaleza, Ceará, Brazil; Graduate Program in Public Health, Ceará State University, Fortaleza, Ceará, Brazil; Department of Nursing, Federal University of Espírito Santo, Vitória, Espírito Santo, Brazil

**Keywords:** Women, Breast cancer, Palliative care, Scoping review

## Abstract

**Introduction:** Palliative care is an approach that improves patients’ quality of life and their families when they face problems inherent to a life-threatening illness. In the current scenario where breast cancer ranks first among the most common types of cancer in women, research has revealed little control by these patients over the symptoms of the disease and its treatment, resulting in poor quality of life, which should be the focus of attention of palliative care. Thus, the purpose of this study is to map and synthesize the available evidence on palliative care in treating breast cancer women.

**Methods:** This scoping review protocol was elaborated following the methodological recommendations proposed by the Joanna Briggs Institute (JBI) and the PRISMA Extension for Scoping Reviews (PRISMA-ScR) checklist. All study designs, experimental studies, observational studies, qualitative studies, mixed studies, and reviews reporting the palliative care for breast cancer women, will be considered. The following electronic databases will be searched: MEDLINE (via PubMed), Embase, Cochrane Library, Web of Science, Scopus, JBI Evidence Synthesis, Epistemonikos, CINAHL (via EBSCO), LILACS, and the electronic repository SciELO. In addition, The British Library, Google Scholar, Preprints for Health Sciences [medRXiv], Open Grey, Who Library Database, ProQuest Global Dissertations and Theses, ClinicalTrials.gov, and the WHO International Clinical Trials Registry Platform will be searched. No language or date restriction will be applied in the search strategy. Two investigators will independently select studies and perform data extraction and critical appraisal using the using JBI design-specific tolos. A narrative synthesis of the evidence will be carried out and will be grouped and presented in tables and graphic models. Also, a similarity analysis will be performed using IRaMuTeQ software version 0.7 alpha 2, leading to textual categories and themes.

**Expected Results:** The findings of this review will help to identify the gaps for the design of future primary research on women with breast cancer in palliative care. Ultimately, will position care services, managers, and health professionals in the knowledge of the phenomenon, so that they can implement the best evidence-based palliative care practices and improve the quality of care provided to breast cancer patients.

**Open Science Framework Registration:** osf.io/5bwq7

## Introduction

Talking about palliative care (PC) today is an interesting challenge due to the complex heterogeneity of conceptions that emerge from this term. However, PC is not a new concept. Saunders [1], the forerunner of the “modern philosophy of hospice,” already emphasized in 1978 the importance of PC in the care of patients with serious illnesses, a premise that today, with the increase in life expectancy and scientific progress of medical sciences has become more relevant. The growing incidence-prevalence of chronic-degenerative diseases has prolonged the “death process”, making it essential to integrate PC into care process towards the end of life.

It is stand out that only 14% of people who need palliative care receive it worldwide. It is estimated that, annually, 40 million people need PC; 78% of them live in low- and middle-income countries that lack training and awareness of PC among health professionals, a major obstacle in addition to the lack of public and government policies aimed at the implementation of PC at the different levels of health care [2].

Palliative care aims to improve the quality of life (QoL) of patients and their families when facing life-threatening situations, and it is essential to improve the well-being, comfort, and dignity of these individuals [3]. In this sense, PC becomes necessary in the therapeutic approach of people with chronic diseases and, despite the negative or passive connotation of the term, palliative treatment must be eminently active, for example, in patients with advanced cancer, who need surgical and radiotherapy treatments to achieve cancer symptom clusters control [4–6].

Worldwide, an estimated 19.3 million new cancer cases (18.1 million excluding nonmelanoma skin cancer) and almost 10.0 million cancer deaths (9.9 million excluding nonmelanoma skin cancer) occurred in 2020 [7]. Female breast cancer has surpassed lung cancer as the most commonly diagnosed cancer, with an estimated 2.3 million new cases (11.7%) [7]. Thus, breast cancer may be of interest for the development of research in the areas of diagnosis, treatment, disease control, and palliative care.

The diagnosis of breast cancer harms the life of women, who experience feelings of fear and suffering throughout the entire process, from diagnosis to therapeutic and survival stages [8,9]. In this context, PC aims to improve chronic disease patients’ and families’ QoL, helping alleviate symptoms and maintain disease control, thus achieving a balance between progression and the discomfort caused by the medical treatment [10].

Studies conducted by Kokkone et al. [11] and the updated Clinical Practice Guideline of the American Society of Clinical Oncology [12] recommend that inpatient and outpatient patients with advanced cancer should receive specific palliative care services, early in the course of the disease, at the same time as active treatment. However, this recommendation is not always met, especially in developing countries with an unmet need for PC [13,14].

Despite the strong degree of recommendation to include PC from diagnosis, recent studies evaluating the QoL in women with breast cancer [8,15–20] have shown a high prevalence of symptoms such as pain, anxiety, fear, cancer-related fatigue, insomnia, dyspnea, and sexual dysfunction, among others. This leads us to think about the current role that PC plays in the treatment of breast cancer, how they are being addressed in this population and their impact on these women’s lives.

In addition, it is important to know the role that PC plays in the healthcare of vulnerable populations, in line with the third Objective of Sustainable Development [21]: “To enable everyone to live in good health and promote the well-being of all at all ages”. Hence, the purpose of this scoping review is to map and synthesize the available evidence on palliative care in treating breast cancer women. The following specific will guide the review:

- What is the scope of the evidence on guidelines or protocols that direct the delivery of palliative care in women with breast cancer?
- What is the extent of the literature on the QoL benefits for women with breast cancer receiving palliative care?

## Materials and methods

### Design

This is a scoping review following the methodological recommendations proposed by the Jonna Briggs Institute (JBI) [22] based on the initial definition of the method proposed by Arskey & O’Malley [23] (Figure 1) as well as the PRISMA Extension for Scoping Reviews (PRISMA-ScR) checklist [24]. The protocol was registered in Open Science Framework (Registration number: osf.io/5bwq7).

**Figure 1.**
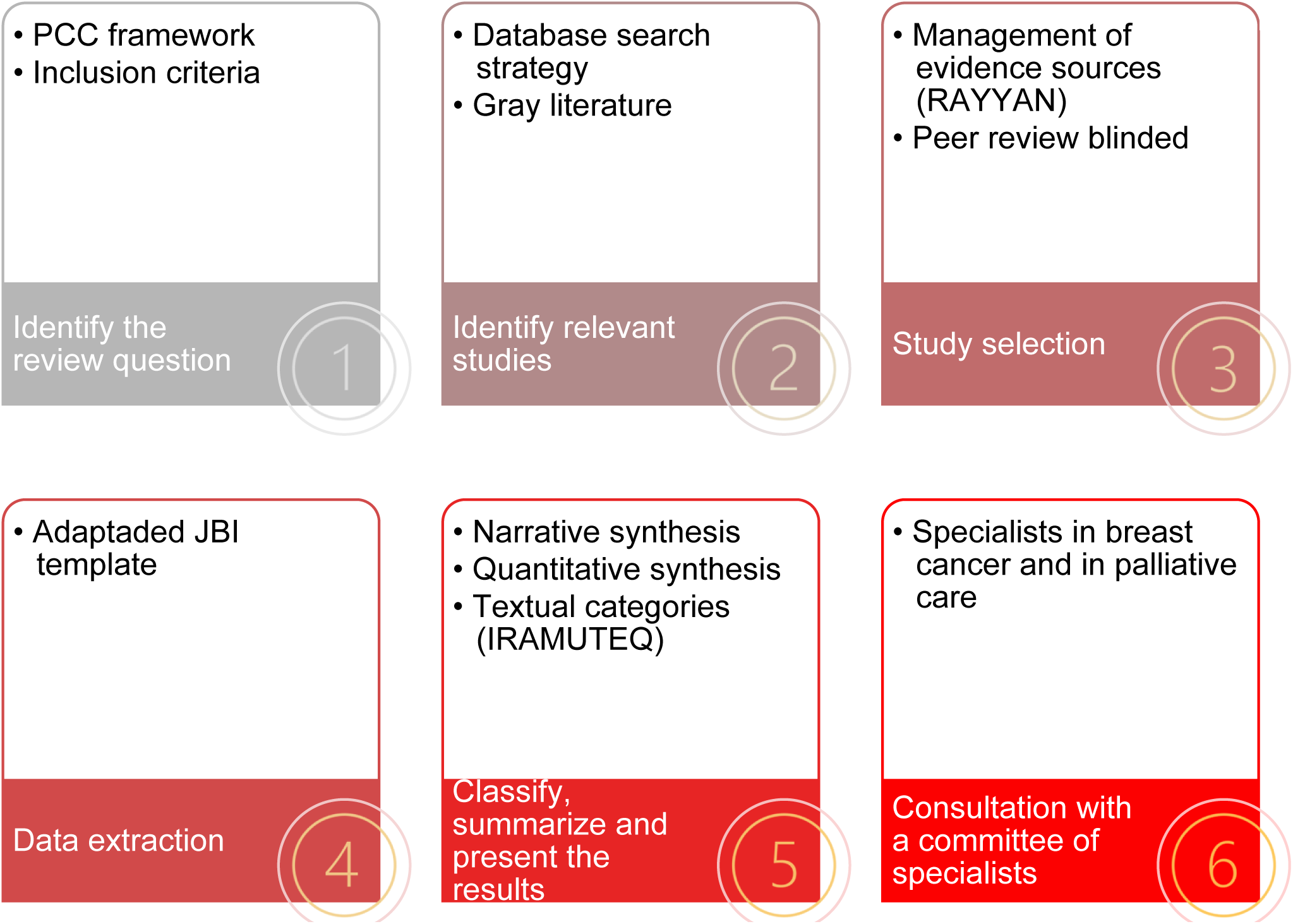
Steps of the scoping review methodology

### Literature review question

The Population, Concept, Context (PCC) framework proposed by the Jonna Briggs Institute (JBI) was used for the construction of the research question [22] as follows: P – young, adult, and older women, C – palliative care, and C – breast cancer, resulting in the following research question: What is the available evidence on palliative care in the treatment of women with breast cancer?

### Inclusion criteria

All studies addressing the variables of the PCC acronym will be included, without restricting the methodological design (e.g., experimental, quasi-experimental, prospective and retrospective cohort, case-control, cross-sectional, case series, individual case reports, descriptive, qualitative, mixed studies, reviews), and no language or publication date restriction. (Figure 2).

**Figure 2.**
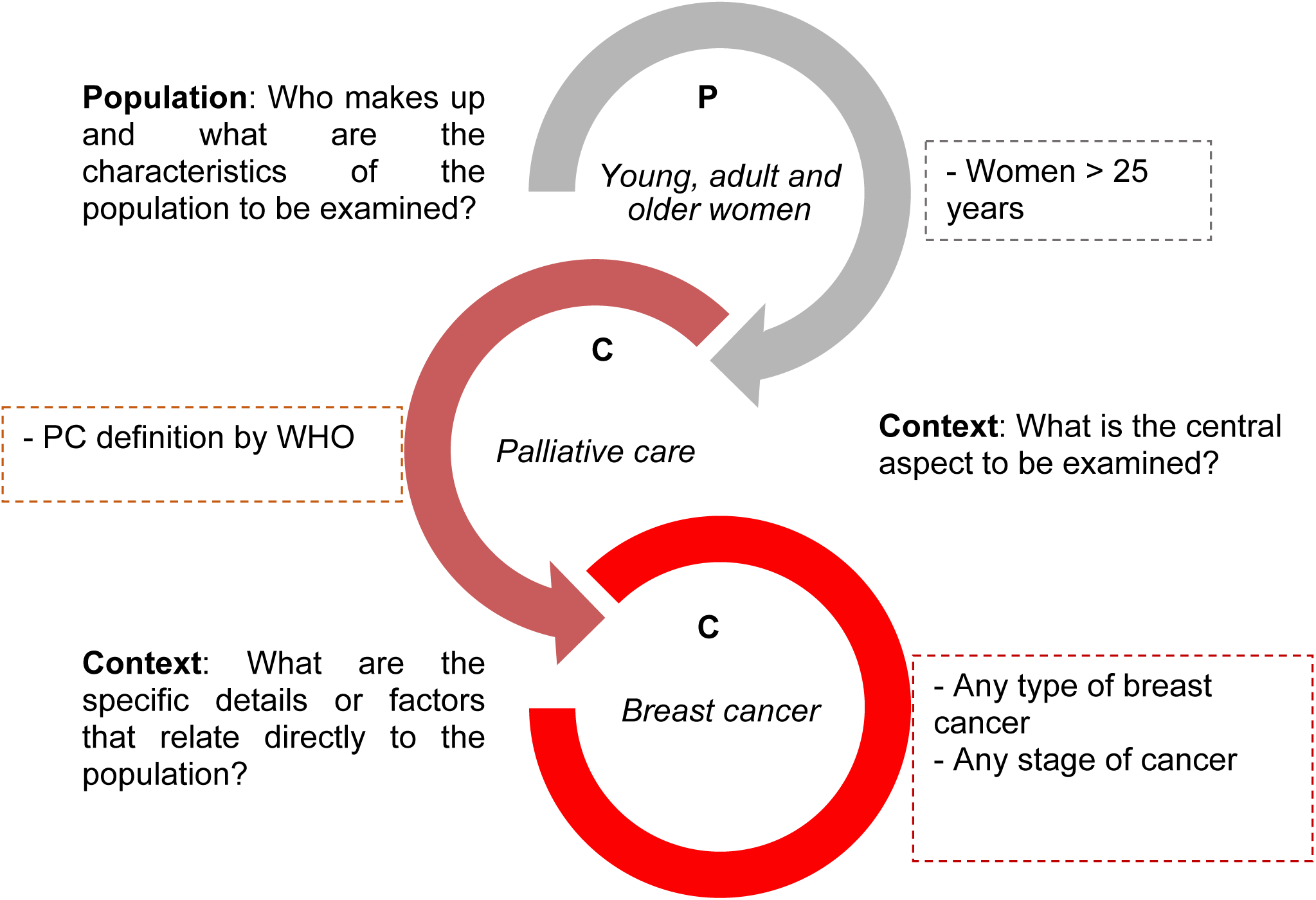
Inclusion criteria according to acronym PCC.

#### Participants

All studies addressing women, from a young age, will be included, according to the age classification proposed by the WHO: “young age: 25 to 44 years, middle age: 44 to 60 years, elderly age: 60 to 75 years, senile age: 75-90, and long-livers: after 90” [25]. This distinction was made because the highest incidence of breast cancer occurs in women ≥ 40 years (26).

#### Concept

Studies in line with the referential framework of palliative care described by the WHO will be included [2], defined as “an approach that improves the quality of life of patients and their families facing the problems associated with life-threatening illness through the prevention and relief of suffering by means of early identification, assessment, and treatment of pain and other problems, physical, psychosocial, and spiritual”.

#### Context

All studies addressing breast cancer will be included, without restricting the type of cancer or stage, according to the definition proposed by the WHO [27]. “Breast cancer arises in the lining cells (epithelium) of the ducts (85%) or lobules (15%) in the glandular tissue of the breast. Initially, the cancerous growth is confined to the duct or lobule (“in situ”), where it generally causes no symptoms and has minimal potential for spread (metastasis). Over time, these in situ (stage 0) cancers may progress and invade the surrounding breast tissue (invasive breast cancer) then spread to the nearby lymph nodes (regional metastasis) or to other organs in the body (distant metastasis)”.

### Exclusion criteria

Studies whose participants have breast cancer diagnosed with other pathologies will be excluded. In addition, studies with male patients with breast cancer will also be excluded.

### Search strategy

The search strategy will be carried out systematically in eight electronic databases: MEDLINE (Via PubMed), Embase (Excerpta Medica Database), LILACS (Latin American and Caribbean Health Sciences Literature), Cochrane Library, Web of Science, Scopus, JBI Evidence Synthesis, Epistemonikos, CINAHL (Cumulative Index to Nursing and Allied Health Literature via EBSCO), as well as the electronic repository SciELO (Scientific Electronic Library Online). It is emphasized that there will be no date or language restrictions in the search strategy. In addition to the electronic databases mentioned above, other sources will be also searshed including: The British Library, Google Scholar, Preprints for Health Sciences [medRXiv], Open Grey, Who Library Database, ProQuest Global Dissertations, and Theses, ClinicalTrials.gov, and the WHO International Clinical Trials Registry Platform.

Due to the flexibility of including gray literature in scoping reviews [22] and the importance of knowing the role of PC in breast cancer, gray literature will also be included [28], such as clinical practice guidelines, protocols, and information contained in the websites of recognized scientific societies that deal with breast cancer and PC.

In addition, the final reference list from the included primary studies will be manually scanned to find relevant studies. Two researchers will search independently, according to the JBI guidelines [22]. Initially, we will identify the existence of an index of specific subject in each database (with MeSH terms, Emtree, DeCS, and CINAHL headings) and their synonyms (keywords). The search terms will then be combined using the Boolean operators “AND” and “OR” [29,30].

The pilot search strategy elaborated with the help and validation of a librarian combining controlled MeSH descriptors and keywords used in MEDLINE/PubMed is as follows:

P- POPULATION:

#1 ((Women) [MeSH Terms] OR Woman [All Fields] OR Female [MeSH Terms] OR Females [All Fields]))

C- CONCEPT: palliative care

#2 ((Palliative care [MeSH Terms] OR Care, palliative [All Fields] OR Palliative treatment [All Fields] OR Palliative treatments [All Fields] OR Treatment palliative [All Fields] OR Treatments palliative [All Fields] OR Therapy palliative [All Fields] OR Palliative therapy [All Fields] OR Palliative Medicine [MeSH Terms] OR Palliative care medicine [All Fields] OR Palliative supportive care [All Fields] OR Supportive care, palliative [All Fields] OR Supportive care [All Fields] OR Palliative surgery [All Fields] OR Surgery, palliative [All Fields] OR Cancer palliative therapy [All Fields]))

#1 AND #2

C- CONCEPT:

#3 ((Breast neoplasms [MeSH Terms] OR Breast neoplasm [All Field] OR Breast tumors [All Fields] OR Breast tumor [All Fields] OR Breast cáncer [All Fields] OR Mammary cancer [MeSH Terms] OR Cancer of breast [All Fields] OR Cancer of the breast [All Fields] OR Mammary carcinoma, human [All Fields]] OR Human mammary carcinomas [All Fields] OR Human mammary carcinoma [All Fields] OR Human mammary neoplasm [All Fields] OR Human mammary neoplasms [MeSH Terms] OR Breast carcinoma [All Fields] OR Breast carcinomas [All Fields] OR Breast malignant neoplasm [All Fields] OR Malignant tumor of breast [All Fields] OR Breast malignant tumors [All Fields] OR Advanced breast cancer [All Fields]))

#3 #1 AND #2

### Selection of the sources of evidence

Rayyan Systems Inc. (RAYYAN) software will use in order to manage the sources of evidence [31]. Initially, all the articles from the general search will be deposited in the RAYYAN software. Then, the duplicates will be removed, and a representative sample of the articles will be taken to evaluate the level of concordance and report the degree of agreement between the two reviewers. Cohen’s kappa coefficient will be used to estimate the index of agreement between the 2 evaluators in each review phase [32].

After this phase, the articles will be assessed by titles and abstracts by two independent researchers to identify potentially eligible articles, and another reviewer will resolve any uncertainty regarding the inclusion of a study. Two reviewers will independently examine the full manuscripts that passed the first selection, and a third reviewer will resolve any decision conflict to obtain a definitive list of studies.

### Data charting

The data will be extracted through a form designed by a research team member adapted to the data extraction template proposed by the JBI and considering previously published extraction forms [29,33-36]. The information to be extracted includes a) study identification and objectives; b) study population and baseline characteristics; c) study design; d) sample size; e) results; f) main findings; g) clinical and epidemiological significance; h) conclusions, i) implications, and j) limitations. Two independent reviewers will extract the data from the included studies.

### Methodological appraisal of included studies

The hierarchy of evidence [37] will classify the selected studies. This classification is divided into seven hierarchical levels, as described in Table 1.

**Table 1.** Hierarchy of evidence.

| **Evidence level** | **Study design** |
| --- | --- |
| I | Evidence from systematic reviews or meta-analyses of randomized controlled clinical trials (RCTs) |
| II | Evidence from a well-designed RCT |
| III | Evidence from well-designed controlled clinical trials without randomization |
| IV | Evidence from well-designed case-control, cohort, or cross-sectional studies |
| V | Evidence from systematic reviews of qualitative and descriptive studies |
| VI | Evidence from a single descriptive or qualitative study |
| VII | Evidence from the opinion of authorities or reports of expert committees |

Two independent reviewers will assess the methodological quality of the studies using JBI design-specific tools [38]. Disagreements will be addressed by the third reviewer.

### Data analysis and Synthesis of results

A flowchart diagram (Figure 3) will describe the entire study selection process [39].

**Figure 3.**
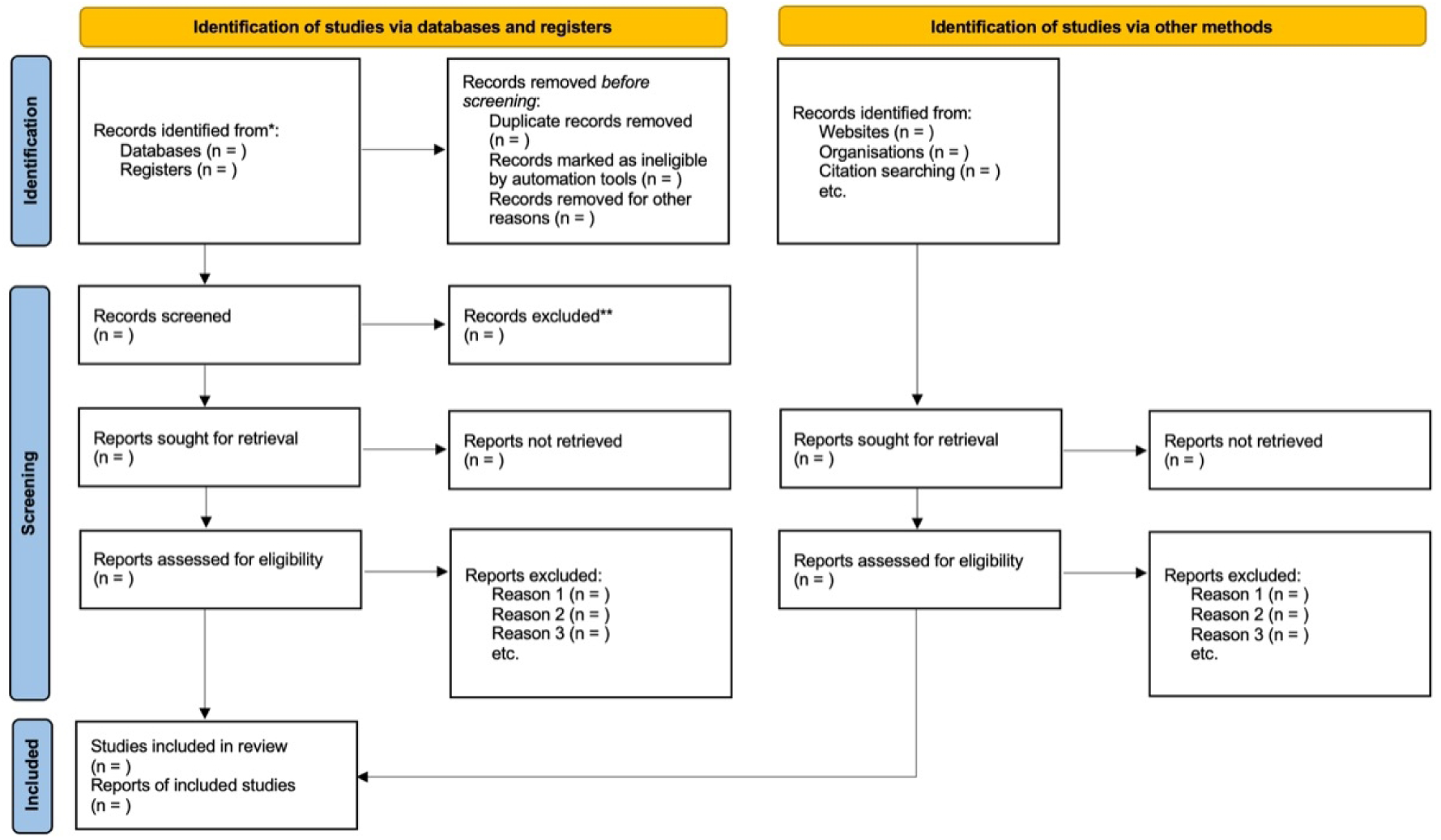
PRISMA flowchart.

A narrative synthesis of the evidence will be carried out, and the primary quantitative data of the included studies will be tabulated. The results will be grouped and presented in tables, and graphic models. Similarity analysis obtained using the IRaMuTeQ software version 0.7 alpha 2 (Interface de R pour les Analyses Multidimensionnelles de Textes et de Questionnaires) will be carried out through textual categories, as appropriate, to define the final themes.

## Discussion and dissemination

Palliative care is essential in treating women with breast cancer, not only because it comprehensively addresses their symptoms but also because it improves their quality of life, and its delivery must be initiated from the diagnosis of the disease. The findings of this review will help to identify the gaps for the design of future research on PC in breast cancer. Ultimately, will position care services, managers, and health professionals in the knowledge of the phenomenon, so that they can implement the best evidence-based PC, consequently improving the quality of care provided to breast cancer patients.

As strong points, this scoping review follows all the methodological recommendations proposed by the JBI. An extensive literature search will be conducted in 9 databases 1 electronic repository. A librarian will guide search strategies, and the final version of this protocol was reviewed and approved by a group of 3 specialists: a breast cancer specialist, a PC specialist, and a scoping review specialist.

## Data Availability

Lopes-Júnior LC, Yanez RJV, Fernandes AFC, Mattos SM, Moreira TMM, Castro RCMB. Palliative care in the treatment of women with breast cancer: a scoping review protocol. 2022. Open Science Framework Repository. https://doi.org/10.17605/OSF.IO/5BWQ7

## Acknowledgements

None to declare.

## References

1. Saunders C. The philosophy of terminal care. The management of terminal malignant disease. 1984;2:232–41.

2. Organización Mundial de la Salud. Cuidados paliativos. Cuidados Paliativos. 2020. Disponível em: https://www.who.int/es/news-room/fact-sheets/detail/palliative-care

3. Organización Mundial de la Salud, Organización Panamericana de la Salud. Cuidados Paliativos [Internet]. Pan American Health Organization / World Health Organization. 2016. Disponível em: https://www3.paho.org/hq/index.php?option=com_content&view=article&id=12587:palliative-care&Itemid=42139&lang=es

4. Instituto Nacional de Câncer. Cuidados paliativos. INCA - Instituto Nacional de Câncer. 2018. Disponível em: https://www.inca.gov.br/controle-do-cancer-do-colo-do-utero/acoes-de-controle/cuidados-paliativos

5. Lopes-Júnior LC, Rosa GS, Pessanha RM, Schuab SIPC, Nunes KZ, Amorim MHC. Efficacy of the complementary therapies in the management of cancer pain in palliative care: A systematic review. Rev Lat Am Enfermagem. 2020 Sep 30;28:e3377. doi: 10.1590/1518-8345.4213.3377.

6. Lopes-Júnior LC, Tuma MC, Amorim MHC. Psychoneuroimmunology and oncology nursing: a theoretical study. Rev Esc Enferm USP. 2021 Sep 10;55:e20210159.

7. Sung H, Ferlay J, Siegel RL, Laversanne M, Soerjomataram I, Jemal A, Bray F. Global Cancer Statistics 2020: GLOBOCAN Estimates of Incidence and Mortality Worldwide for 36 Cancers in 185 Countries. CA Cancer J Clin. 2021 May;71(3):209–249. doi: 10.3322/caac.21660.

8. Lopes JV, Bergerot CD, Barbosa LR, Calux NMCT, Elias S, Ashing KT, Domenico EBL. Impact of breast cancer and quality of life of women survivors. Rev Bras Enferm. 2018 Nov-Dec;71(6):2916–2921. doi: 10.1590/0034-7167-2018-0081.

9. Abrahão CA, Bomfim E, Lopes-Júnior LC, Pereira-da-Silva G. Complementary Therapies as a Strategy to Reduce Stress and Stimulate Immunity of Women With Breast Cancer. J Evid Based Integr Med. 2019 Jan-Dec;24:2515690X19834169. doi: 10.1177/2515690X19834169.

10. Drageset S, Austrheim G, Ellingsen S. Quality of life of women living with metastatic breast cancer and receiving palliative care: A systematic review. Health Care Women Int. 2021 Sep;42(7-9):1044-1065. doi: 10.1080/07399332.2021.1876063.

11. Kokkonen K, Tasmuth T, Lehto JT, Kautiainen H, Elme A, Jääskeläinen AS, Saarto T. Cancer Patients’ Symptom Burden and Health-related Quality of Life (HRQoL) at Tertiary Cancer Center from 2006 to 2013: A Cross-sectional Study. Anticancer Res. 2019 Jan;39(1):271–277. doi: 10.21873/anticanres.13107.

12. Ferrell BR, Temel JS, Temin S, Alesi ER, Balboni TA, Basch EM, et al. Integration of Palliative Care Into Standard Oncology Care: American Society of Clinical Oncology Clinical Practice Guideline Update. JCO 2017;35(1):96–112. Disponível em: https://ascopubs.org/doi/10.1200/JCO.2016.70.1474

13. Reid EA, Gudina EK, Ayers N, Tigineh W, Azmera YM. Caring for Life-Limiting Illness in Ethiopia: A Mixed-Methods Assessment of Outpatient Palliative Care Needs. J Palliat Med. 2018 May;21(5):622–630. doi: 10.1089/jpm.2017.0419.

14. Kaba M, de Fouw M, Deribe KS, Abathun E, Peters AAW, Beltman JJ. Palliative care needs and preferences of female patients and their caregivers in Ethiopia: A rapid program evaluation in Addis Ababa and Sidama zone. PLoS One. 2021 Apr 22;16(4):e0248738. doi: 10.1371/journal.pone.0248738.

15. Haddou Rahou B, El Rhazi K, Ouasmani F, Nejjari C, Bekkali R, Montazeri A, Mesfioui A. Quality of life in Arab women with breast cancer: a review of the literature. Health Qual Life Outcomes. 2016 Apr 27;14:64. doi: 10.1186/s12955-016-0468-9.

16. Villar RR, Fernández SP, Garea CC, Pillado MTS, Barreiro VB, Martín CG. Quality of life and anxiety in women with breast cancer before and after treatment. Rev Lat Am Enfermagem. 2017 Dec 21;25:e2958. doi: 10.1590/1518-8345.2258.2958.

17. Ghislain I, Zikos E, Coens C, Quinten C, Balta V, Tryfonidis K, Piccart M, Zardavas D, Nagele E, Bjelic-Radisic V, Cardoso F, Sprangers MAG, Velikova G, Bottomley A; European Organisation for Research and Treatment of Cancer (EORTC) Quality of Life Group; Breast Cancer Group; EORTC Headquarters. Health-related quality of life in locally advanced and metastatic breast cancer: methodological and clinical issues in randomised controlled trials. Lancet Oncol. 2016 Jul;17(7):e294–e304. doi: 10.1016/S1470-2045(16)30099-7.

18. Mokhtari-Hessari P, Montazeri A. Health-related quality of life in breast cancer patients: review of reviews from 2008 to 2018. Health Qual Life Outcomes. 2020 Oct 12;18(1):338. doi: 10.1186/s12955-020-01591-x.

19. Salibasic M, Delibegovic S. The Quality of Life and Degree of Depression of Patients Suffering from Breast Cancer. Med Arch. 2018 Jun;72(3):202–205. doi: 10.5455/medarh.2018.72.202-205.

20. Guerra RL, Dos Reis NB, Corrêa FM, Fernandes MM, Ribeiro Alves Fernandes R, Cancela MC, Araújo RM, Crocamo S, Santos M, De Almeida LM. Breast Cancer Quality of Life and Health-state Utility at a Brazilian Reference Public Cancer Center. Expert Rev Pharmacoecon Outcomes Res. 2020 Apr;20(2):185–191. doi: 10.1080/14737167.2019.1621752.

21. Organización Mundial de la Salud. Objetivo 3: Garantizar una vida sana y promover el bienestar para todos en todas las edades. Objetivos delDesarrollo Sostenible. 2020. Disponível em: https://www.un.org/sustainabledevelopment/es/health/

22. Aromataris E, Munn Z, Org. JBI Manual for Evidence Synthesis [Internet]. JBI; 2020. Disponível em: https://wiki.jbi.global/display/MANUAL

23. Levac D, Colquhoun H, O’Brien KK. Scoping studies: advancing the methodology. Implement Sci. 2010 Sep 20;5:69. doi: 10.1186/1748-5908-5-69. PMID: 20854677; PMCID: PMC2954944.

24. Tricco AC, Lillie E, Zarin W, O’Brien KK, Colquhoun H, Levac D, Moher D, Peters MDJ, Horsley T, Weeks L, Hempel S, Akl EA, Chang C, McGowan J, Stewart L, Hartling L, Aldcroft A, Wilson MG, Garritty C, Lewin S, Godfrey CM, Macdonald MT, Langlois EV, Soares-Weiser K, Moriarty J, Clifford T, Tunçalp Ö, Straus SE. PRISMA Extension for Scoping Reviews (PRISMA-ScR): Checklist and Explanation. Ann Intern Med. 2018 Oct 2;169(7):467–473. doi: 10.7326/M18-0850.

25. Zhaoyang R, Sliwinski MJ, Martire LM, Smyth JM. Age differences in adults’ daily social interactions: An ecological momentary assessment study. Psychol Aging. 2018 Jun;33(4):607–618. doi: 10.1037/pag0000242. Epub 2018 Apr 30. PMID: 29708385; PMCID: PMC6113687.

26. Coughlin SS. Epidemiology of Breast Cancer in Women. In: Ahmad A, organizador. Breast Cancer Metastasis and Drug Resistance: Challenges and Progress. Cham: Springer International Publishing; 2019:9–29. Disponível em: https://doi.org/10.1007/978-3-030-20301-6_2

27. Organización Mundial de la Salud. Cáncer de mama. Cáncer de Mama. 2021. Disponível em: https://www.who.int/es/news-room/fact-sheets/detail/breast-cancer

28. Botelho RG, Oliveira C da C de. Literaturas branca e cinzenta: uma revisão conceitual. Ciência da Informação. 2015;44(3). Disponível em: http://revista.ibict.br/ciinf/article/view/1804

29. Lopes-Júnior LC, Siqueira PC, Maciel ELN. School reopening and risks accelerating the COVID-19 pandemic: A systematic review and meta-analysis protocol. PLoS One. 2021 Nov 17;16(11):e0260189. doi: 10.1371/journal.pone.0260189.

30. Gonçalves CA, Lopes-Júnior LC, Nampo FK, Zilly A, Mayer PCM, Pereira-da-Silva G. Safety, efficacy and immunogenicity of therapeutic vaccines in the treatment of patients with high-grade cervical intraepithelial neoplasia associated with human papillomavirus: a systematic review protocol. BMJ Open. 2019 Jul 17;9(7):e026975. doi: 10.1136/bmjopen-2018-026975.

31. Ouzzani M, Hammady H, Fedorowicz Z, Elmagarmid A. Rayyan-a web and mobile app for systematic reviews. Syst Rev. 2016 Dec 5;5(1):210. doi: 10.1186/s13643-016-0384-4.

32. McHugh ML. Interrater reliability: the kappa statistic. Biochem Med (Zagreb). 2012;22(3):276–82. PMID: 23092060; PMCID: PMC3900052.

33. Lopes-Júnior LC, Bomfim E, Olson K, Neves ET, Silveira DSC, Nunes MDR, Nascimento LC, Pereira-da-Silva G, Lima RAG. Effectiveness of hospital clowns for symptom management in paediatrics: systematic review of randomised and non-randomised controlled trials. BMJ. 2020 Dec 16;371:m4290. doi: 10.1136/bmj.m4290.

34. Lopes-Júnior LC, Bomfim EO, Nascimento LC, Nunes MD, Pereira-da-Silva G, Lima RA. Non-pharmacological interventions to manage fatigue and psychological stress in children and adolescents with cancer: an integrative review. Eur J Cancer Care (Engl). 2016 Nov;25(6):921–935. doi: 10.1111/ecc.12381.

35. Armstrong R, Hall BJ, Doyle J, Waters E. Cochrane Update. ’Scoping the scope’ of a cochrane review. J Public Health (Oxf). 2011 Mar;33(1):147–50. doi: 10.1093/pubmed/fdr015. PMID: 21345890.

36. Valaitis R, Martin-Misener R, Wong ST, MacDonald M, Meagher-Stewart D, Austin P, Kaczorowski J, O-Mara L, Savage R; Strengthening Primary Health Care through Public Health and Primary Care Collaboration Team. Methods, strategies and technologies used to conduct a scoping literature review of collaboration between primary care and public health. Prim Health Care Res Dev. 2012 Jul;13(3):219–36. doi: 10.1017/S1463423611000594.

37. Fineout-Overholt E, Melnyk BM, Stillwell SB, Williamson KM. Evidence-based practice step by step: Critical appraisal of the evidence: part I. Am J Nurs. 2010 Jul;110(7):47–52. doi: 10.1097/01.NAJ.0000383935.22721.9c.38.

38. Critical Appraisal Tools - JBI. 2017. Disponível em http://joannabriggs.org/research/critical-appraisal-tools.html

39. Page MJ, McKenzie JE, Bossuyt PM, Boutron I, Hoffmann TC, Mulrow CD, et al. The PRISMA 2020 statement: an updated guideline for reporting systematic reviews. BMJ. 2021 Mar 29;372:n71. doi: 10.1136/bmj.n71.

